# The COvid-19 Pandemic and Exercise (COPE) Trial: A multi-group randomized controlled trial comparing effects of an app-based, at-home exercise program to waitlist control on depressive symptoms

**DOI:** 10.1101/2021.04.14.21255519

**Authors:** Eli Puterman, Benjamin A. Hives, Nicole Mazara, Nikol Grishin, Joshua Webster, Stacey Hutton, Michael Koehle, Yan Liu, Mark R Beauchamp

## Abstract

**Background:** The number of adults across the globe with significant depressive symptoms has grown substantially during the COVID-19 pandemic. The extant literature supports exercise as a potent behavior that can significantly reduce depressive symptoms in clinical and non-clinical populations.

**Objective:** Using a suite of mobile applications, at-home exercise, including high intensity interval training (HIIT) and/or yoga, was completed to reduce depressive symptoms in the general population in the early months of the pandemic.

**Methods:** A 6-week, parallel, multi-arm, randomized controlled trial was completed with 4 groups: [1] HIIT, [2] Yoga, [3] HIIT+Yoga, and [4] waitlist control (WLC). Low active, English-speaking, non-retired Canadians aged 18-64 years were included. Depressive symptoms were measured at baseline and weekly following randomization.

**Results:** A total of 334 participants were randomized to one of four groups. No differences in depressive symptoms were evident at baseline. The results of latent growth modeling showed significant treatment effects for each active group compared to the WLC, with small effect sizes in the community-based sample of participants. Treatment groups were not significantly different from each other. Effect sizes were very large when restricting analyses only to participants with high depressive symptoms at baseline.

**Conclusions:** At-home exercise is a potent behavior to improve mental health in adults during the pandemic, especially in those with increased levels of depressive symptoms. Promotion of at-home exercise may be a global public health target with important personal, social, and economic implications as the world emerges scathed by the pandemic.

**Trial registration number:** clinicaltrials.gov #NCT04400279

**Summary Box:** This randomized controlled trial provides strong evidence suggesting that at-home app-based exercise in various forms (high intensity interval training or yoga or their combination) can significantly improve depression symptoms over a 6-week period in community adults during the pandemic. When the sample was restricted to only those with high baseline depression symptoms, the weekly effects were substantially large. At-home exercise during the COVID-19 pandemic proved to be an impactful and affordable health behavior in which community living adults, especially those with high depression symptoms, can engage to bolster their mental health. In light of the long-term mental health consequences of COVID-19 with which many adults are expected to struggle, even after a return to normal, promoting and supporting programming in communities at the individual level will emerge as a necessary health policy initiative.

## Introduction

Early in the COVID-19 pandemic, fears of infection, economic hardship, and global stay-at-home mandates were widely predicted to have substantial negative effects on mental health.^1–3^ The prediction has been borne out. A nationally representative study in 1,441 United States (US) residents reported that 27.8% of Americans experienced depressive symptoms in April 2020 compared to 8.5% two years earlier, a greater than 3-fold increase.^4^ Declines in mental health during the early months of the pandemic from pre-pandemic levels were also apparent in a national survey completed in the United Kingdom (UK).^5^

Researchers and healthcare professionals promoted a wide range of approaches to maintain the mental health of all individuals during this pandemic, from actions individuals can take within their homes and outdoors, such as exercise, to assessment and treatment considerations that healthcare providers and institutions can implement.^1–3^ The World Health Organization^6^ and global government agencies (e.g., US Center for Disease Control^7^ and Public Health England^8^) similarly recommended that the public engage in physical activity and exercise to attain and maintain mental health during the pandemic. These recommendations were supported by an extant literature providing compelling evidence for impactful prevention of^9^ and reductions in^10^ depressive symptoms in clinical and non-clinical populations following the adoption of physical activity programming. Yet, with the mandated closure of fitness centres and outdoor recreation sites (e.g., local and state/provincial parks) at the start of the pandemic, opportunities for engaging in healthy behaviours remained limited to one’s home for the most part.

Our study tested whether completing exercise at home that required little physical space or equipment would lead to reductions in depressive symptoms among community-dwelling adults in the Spring and Summer of 2020. Activities were completed with the use of a mobile application, with memberships provided free. The provision of free memberships and the minimal room or equipment required to complete the activities attended to some of the socioeconomic inequities that can result from at-home exercise during the pandemic, as highlighted by Sallis and colleagues.^11^ We partnered with a mobile application company with a suite of applications for a variety of activities that require little space or equipment, including whole body weight-based high intensity interval training (HIIT) and yoga. Both HIIT^12^ and yoga^13^ have been shown to be effective in improving depressive symptoms. Participants in the active groups received access to either the HIIT or yoga application or access to both applications for 6 weeks.

The primary hypothesis was that completing at-home HIIT and/or yoga with the use of a free mobile application will lead to significant declines in depressive symptoms in adults over a 6-week period compared to a waitlist control (WLC). We further tested whether the benefits were unique to or stronger for HIIT, yoga, or their combination by comparing each group’s effects over time to one another. Finally, we tested whether the effects were more or less apparent in those with high depressive symptoms pre-randomization.

## Methods

### Trial Design

The COvid-19 Pandemic and Exercise (COPE) Trial was a parallel randomized controlled trial, with participants allocated randomly to 1 of 4 treatment groups: 1) HIIT, 2) Yoga, 3) HIIT+Yoga, or 4) WLC. Study protocol was approved by the University of British Columbia’s Behavioral Review Ethics Board (#H20-01497), and registered at clinicaltrials.gov (#NCT04400279) and the Open Science Framework (https://osf.io/jbm63/).

### Participants

Low active, English-speaking, Canadians aged 18-64, who were not retired at study entry and had access to the Internet via a mobile device or computer were eligible to participate. Activity was assessed with the validated Stanford Leisure-Time Categorical Activity Item (L-CAT)^14^ and those who scored between 1 and 3 on the item were eligible to participate as these scores represent low activity as prescribed by the American College of Sports Medicine.^15^ Only those deemed capable of performing moderate intensity physical activity were eligible to participate, as assessed with the Physical Activity Readiness Questionnaire for Everyone (PAR-Q+), and if necessary, further assessed with the Physical Activity Readiness Medical Examination (ePARmed-X+).^16^ Those hospitalized within the previous 3 months were not eligible, unless a note from their physician was provided stating their ability to participate.

Participants from across Canada were recruited through social media advertisements (i.e., Facebook, Twitter, Instagram), that directed adults to the study website (www.copetrial.ca). If interested, they were invited to email the study team to schedule an eligibility screening phone call. A study team member completed the screening using a scripted interview with Qualtrics, where eligibility screening data were stored. Upon agreement, eligible and interested participants received two links by email (1) to the Qualtrics-based consent form for electronic signature and (2) to the PAR-Q+. Those who were not cleared for exercise based on the PAR-Q+ or the ePARmed-X+ were required to receive clearance from their family physician or our team’s study physician. Following clearance, all participants completed our Qualtrics-based survey to assess the primary outcome: depressive symptoms.

### Intervention

Those randomized to one of the 3 treatment groups received a free 3-month membership to the mobile application version from DownDog (www.downdogapp.com) to access the applicable programs according to group assignment. Participants in the treatment groups were asked to complete four weekly 20-minute sessions. The intervention was 6 weeks long, with weekly surveys of depressive symptoms. After 6 weeks, WLC participants received the free 3-month membership to the yoga and HIIT apps. To ensure anonymity on the DownDog platform, each participant received a Participant ID that was pre-registered by a study team member on the DownDog platform. This also allowed us to track their weekly progress.

### Outcome

The primary outcome was depressive symptoms, measured weekly from baseline to the end of the 6^th^ week of the trial with the 10-item Center for Epidemiological Studies – Depression Scale (CESD)^17^. Examples of items include, “I was bothered by things that usually don’t bother me,” and “I could not get going.” Scores ranged between 0 (“Rarely or none of the time (less than 1 day)”) and 3 (“Most or all of the time (5-7 days)”). Sum scores were produced (potential range from 0 to 30 (Sample range: 0 to 30)). A cut-off score of 10 or above is considered significant depressive symptoms in community samples.^18^

### Sample Size

Using Optimal Design Software,^19^ to detect a small effect size (ES) δ = .30 based on a two-level curvilinear growth model with Power (1 - b) set at .80 and alpha set at .05 for a seven time points repeated measures design with four groups, 367 participants were required. A 25% attrition was expected over 6 weeks, thus a sample size of 490 was considered to be sufficient for the trial.

### Randomization and Allocation

Sequence generation for randomization was completed using Excel. Each member of the recruiting team received an Excel book with multiple blocks of randomized groupings. All blocks contained one of each potential grouping and a randomly assigned number generated by the data manager (second author). The treatment groups within each block were then sorted by their randomized number, allowing for unique configurations within each block and the grouping hidden on the Excel sheet. A member of the study team would unhide the result of the randomization and allocated the participants to their group once they had completed the baseline surveys. The data manager and PI (first author) remained blind to the participants’ allocations throughout the trial. The PI was blind to all randomization until all data were prepared for analyses and initial primary analysis was completed.

### Changes to the Trial

On August 7^th^, 2020, study recruitment was terminated for several reasons. First, interest in the study dropped substantially in late June, potentially due to the fact that fitness centres across Canada started to open, as did parks. Second, we wanted to keep the time frame of recruitment narrow to maintain similarity across participants in terms of impact of the pandemic. Finally, while we were very conservative with our original expected ES, a small-to-medium ES of δ = .40 or a medium ES of .50 could have as easily been expected based on previous meta-analyses^10,20^, requiring 209 and 134 participants, respectively, or 278 and 179, accounting for 25% attrition. More information on these changes can be accessed here, submitted August 8^th^, 2020: https://osf.io/a65vd/.

### Statistical Approach

Means and standard deviations, or number and percentages, were calculated for all continuous or categorical sociodemographic variables, respectively. ANOVAs or chi-square analyses were completed for the continuous and categorical factors, respectively, to compare group differences. Imputation, using random forest methods,^21^ was conducted for depression symptom score when the participant did not complete all items in the survey at any week (See Supplemental Statistical Approach for information on imputation procedures and survey items completion rates on Table S1). All descriptive statistics, multiple imputation, and visualisations were run using R Statistical software (version 4.0.2).

All randomized participants were included in the intent-to-treat analyses using Mplus (Version 7.2).^22^ We adopted quadratic latent growth models^22^ based on the framework of structural equation models (SEM) to account for non-linear trends in CESD scores over the 6 weeks. First, we conducted an unconditional growth model to estimate intercept (I), slope (S), and quadratic (Q) terms. Next, we included three dummy-coded variables for the active groups, with WLC set as the comparator, to test the prespecified treatment effects of each active group on I, S, and Q. To examine the treatment effects on the subpopulation with high depressive symptoms initially, we restricted the sample to participants with CESD scores of ≥10^17^ and, due to poor model fit when including the quadratic term, we utilized free time scores of the slope growth factor for non-linear trends.^22,p124^ In these analyses, only I and S were estimated. For all analyses, we computed effect sizes at each week using Feingold’s approach,^23^ which is equivalent to Cohen’s *d*. Fit statistics for the analyses with the complete sample and depression-restricted sample are included in the Table S2. All codes are included in the Supplemental MPlus Codes.

## Results

### Participants

Three hundred and ninety-six individuals were screened for the COPE trial and, based on eligibility, 334 (84%) were enrolled between [May 27, 2020] and [August 7, 2020] (see Figure 1, Consort Diagram). Descriptive statistics for the sociodemographic factors and depressive symptoms are presented in Table 1. Treatment groups were not different from each other on any factor at baseline.

**Table 1 –.**
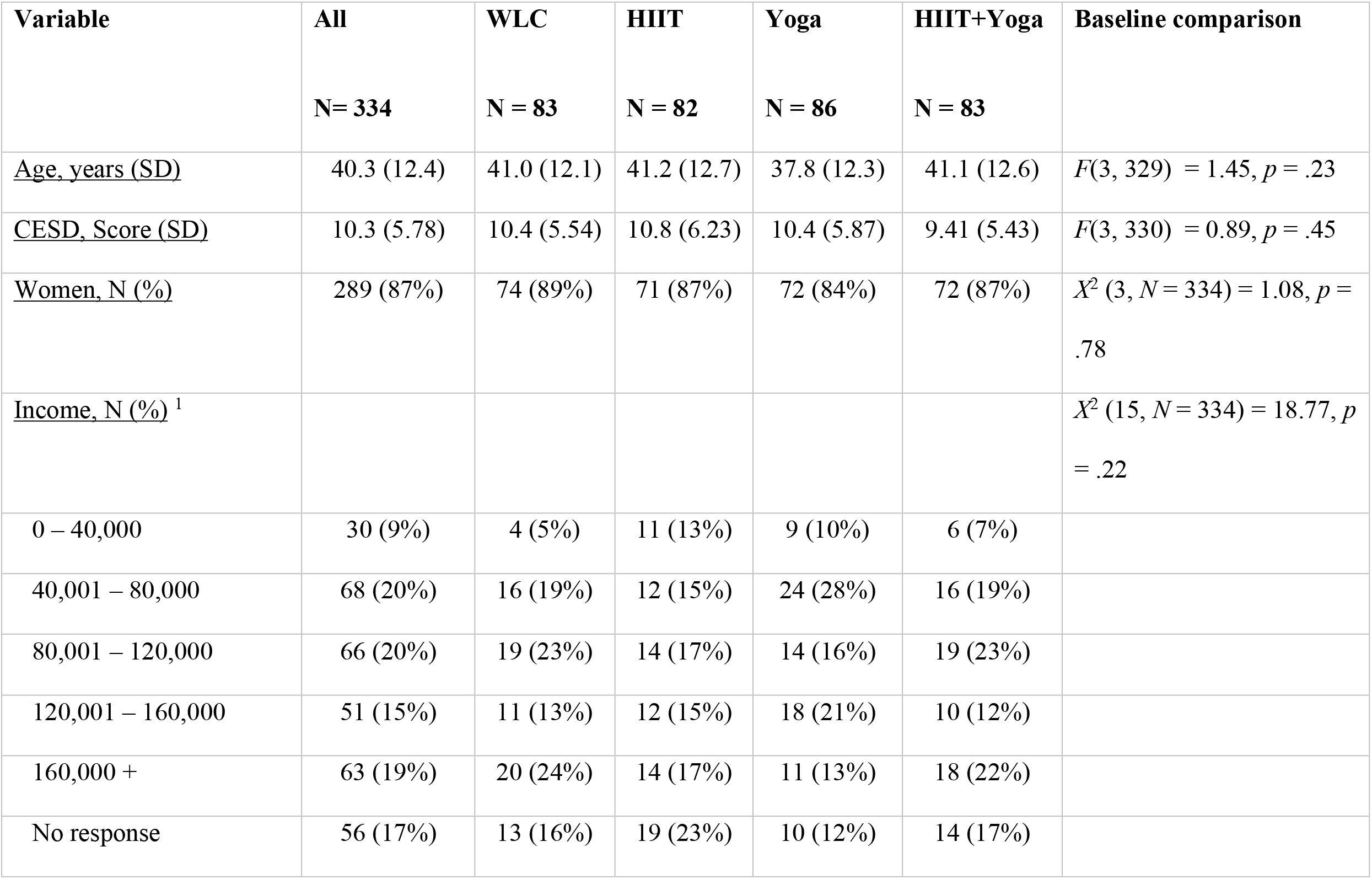
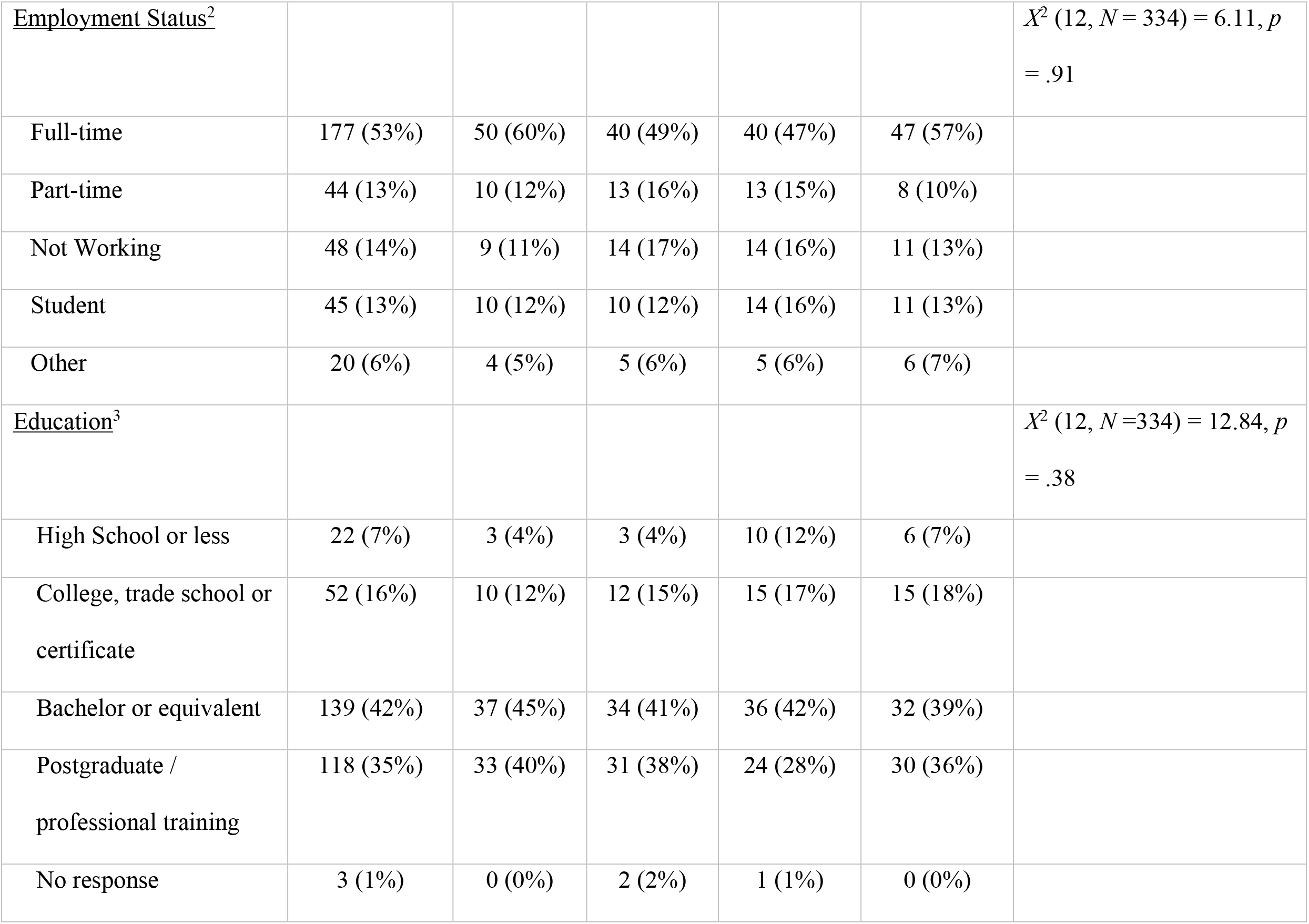
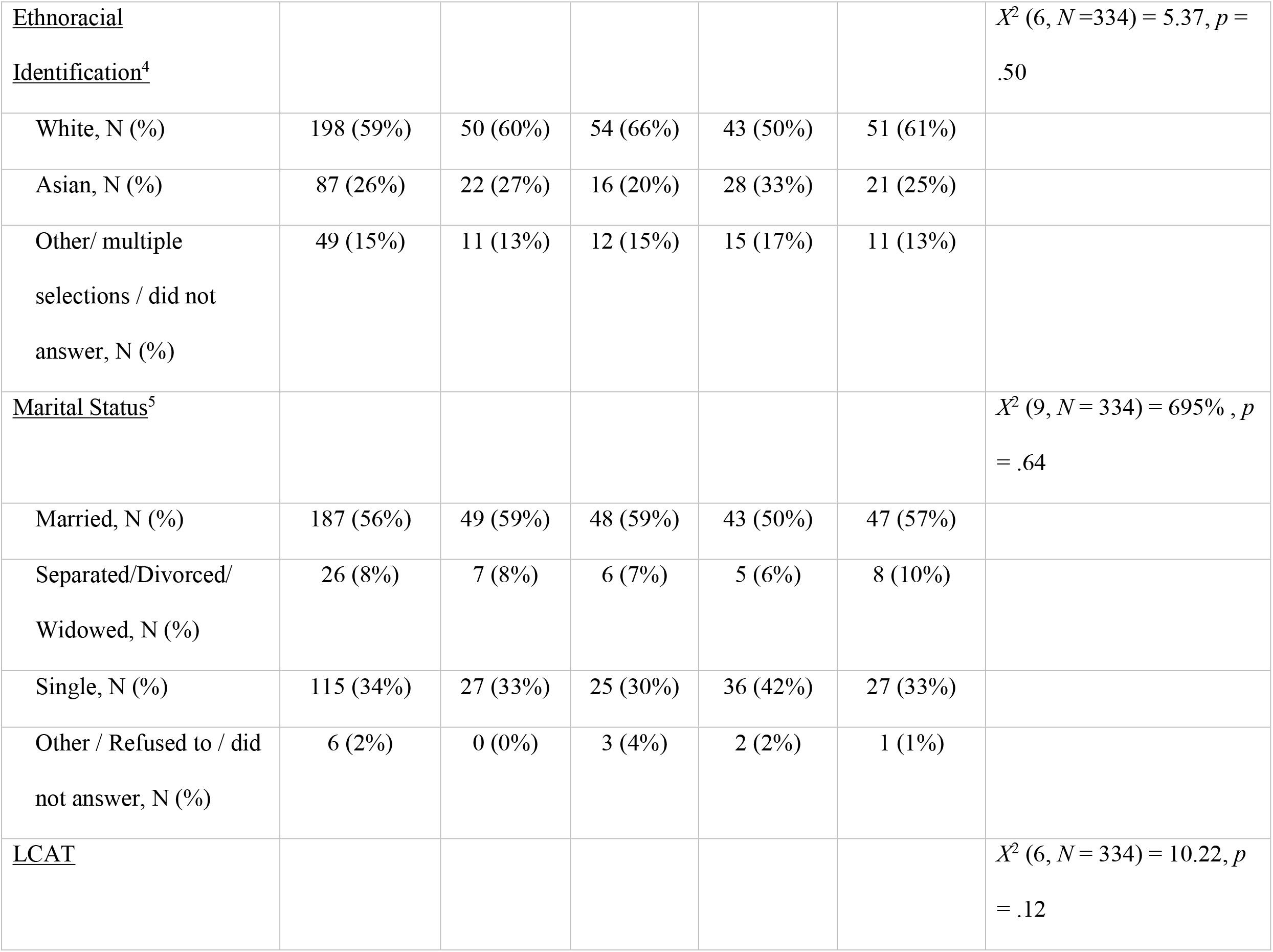
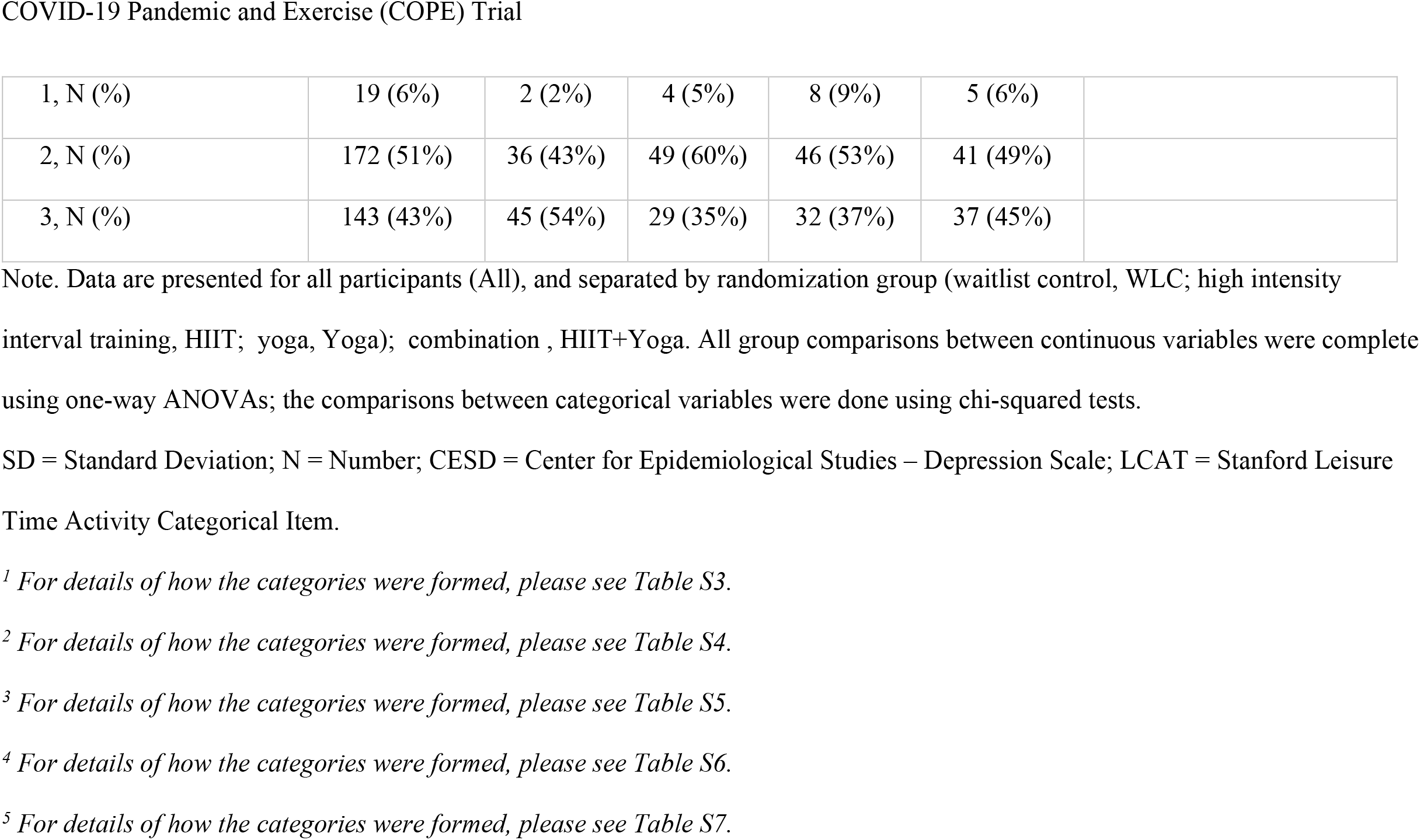
Participant Demographic Information.

| **Variable** | **All** | **WLC** | **HIIT** | **Yoga** | **HIIT+Yoga** | **Baseline comparison** |
| --- | --- | --- | --- | --- | --- | --- |
|  | **N= 334** | **N = 83** | **N = 82** | **N = 86** | **N = 83** |  |
| <u>Age, years (SD)</u> | 40.3 (12.4) | 41.0 (12.1) | 41.2 (12.7) | 37.8 (12.3) | 41.1 (12.6) | $F(3, 329) = 1.45, p = .23$ |
| <u>CESD, Score (SD)</u> | 10.3 (5.78) | 10.4 (5.54) | 10.8 (6.23) | 10.4 (5.87) | 9.41 (5.43) | $F(3, 330) = 0.89, p = .45$ |
| <u>Women, N (%)</u> | 289 (87%) | 74 (89%) | 71 (87%) | 72 (84%) | 72 (87%) | $\chi^2 (3, N = 334) = 1.08, p = .78$ |
| <u>Income, N (%)</u><sup>1</sup> | | | | | | $\chi^2 (15, N = 334) = 18.77, p = .22$ |
| 0 – 40,000 | 30 (9%) | 4 (5%) | 11 (13%) | 9 (10%) | 6 (7%) |  |
| 40,001 – 80,000 | 68 (20%) | 16 (19%) | 12 (15%) | 24 (28%) | 16 (19%) |  |
| 80,001 – 120,000 | 66 (20%) | 19 (23%) | 14 (17%) | 14 (16%) | 19 (23%) |  |
| 120,001 – 160,000 | 51 (15%) | 11 (13%) | 12 (15%) | 18 (21%) | 10 (12%) |  |
| 160,000 + | 63 (19%) | 20 (24%) | 14 (17%) | 11 (13%) | 18 (22%) |  |
| No response | 56 (17%) | 13 (16%) | 19 (23%) | 10 (12%) | 14 (17%) |  |
| <u>Employment Status</u><sup>2</sup> | | | | | | $\chi^2 (12, N = 334) = 6.11, p = .91$ |
| Full-time | 177 (53%) | 50 (60%) | 40 (49%) | 40 (47%) | 47 (57%) |  |
| Part-time | 44 (13%) | 10 (12%) | 13 (16%) | 13 (15%) | 8 (10%) |  |
| Not Working | 48 (14%) | 9 (11%) | 14 (17%) | 14 (16%) | 11 (13%) |  |
| Student | 45 (13%) | 10 (12%) | 10 (12%) | 14 (16%) | 11 (13%) |  |
| Other | 20 (6%) | 4 (5%) | 5 (6%) | 5 (6%) | 6 (7%) |  |
| <u>Education</u><sup>3</sup> | | | | | | $\chi^2 (12, N = 334) = 12.84, p = .38$ |
| High School or less | 22 (7%) | 3 (4%) | 3 (4%) | 10 (12%) | 6 (7%) |  |
| College, trade school or certificate | 52 (16%) | 10 (12%) | 12 (15%) | 15 (17%) | 15 (18%) |  |
| Bachelor or equivalent | 139 (42%) | 37 (45%) | 34 (41%) | 36 (42%) | 32 (39%) |  |
| Postgraduate / professional training | 118 (35%) | 33 (40%) | 31 (38%) | 24 (28%) | 30 (36%) |  |
| No response | 3 (1%) | 0 (0%) | 2 (2%) | 1 (1%) | 0 (0%) |  |
| <u>Ethnoracial Identification<sup>4</sup></u> | | | | | | $\chi^2 (6, N=334) = 5.37, p = .50$ |
| White, N (%) | 198 (59%) | 50 (60%) | 54 (66%) | 43 (50%) | 51 (61%) |  |
| Asian, N (%) | 87 (26%) | 22 (27%) | 16 (20%) | 28 (33%) | 21 (25%) |  |
| Other/ multiple selections / did not answer, N (%) | 49 (15%) | 11 (13%) | 12 (15%) | 15 (17%) | 11 (13%) |  |
| <u>Marital Status<sup>5</sup></u> | | | | | | $\chi^2 (9, N = 334) = 6.95, p = .64$ |
| Married, N (%) | 187 (56%) | 49 (59%) | 48 (59%) | 43 (50%) | 47 (57%) |  |
| Separated/Divorced/<br>Widowed, N (%) | 26 (8%) | 7 (8%) | 6 (7%) | 5 (6%) | 8 (10%) |  |
| Single, N (%) | 115 (34%) | 27 (33%) | 25 (30%) | 36 (42%) | 27 (33%) |  |
| Other / Refused to / did not answer, N (%) | 6 (2%) | 0 (0%) | 3 (4%) | 2 (2%) | 1 (1%) |  |
| <u>LCAT</u> | | | | | | $\chi^2 (6, N = 334) = 10.22, p = .12$ |

|  |  |  |  |  |  |
| --- | --- | --- | --- | --- | --- |
| 1, N (%) | 19 (6%) | 2 (2%) | 4 (5%) | 8 (9%) | 5 (6%) |
| 2, N (%) | 172 (51%) | 36 (43%) | 49 (60%) | 46 (53%) | 41 (49%) |
| 3, N (%) | 143 (43%) | 45 (54%) | 29 (35%) | 32 (37%) | 37 (45%) |

Note. Data are presented for all participants (All), and separated by randomization group (waitlist control, WLC; high intensity interval training, HIIT; yoga, Yoga); combination , HIIT+Yoga. All group comparisons between continuous variables were complete using one-way ANOVAs; the comparisons between categorical variables were done using chi-squared tests.
SD = Standard Deviation; N = Number; CESD = Center for Epidemiological Studies – Depression Scale; LCAT = Stanford Leisure Time Activity Categorical Item.
<sup>1</sup> For details of how the categories were formed, please see Table S3.
<sup>2</sup> For details of how the categories were formed, please see Table S4.
<sup>3</sup> For details of how the categories were formed, please see Table S5.
<sup>4</sup> For details of how the categories were formed, please see Table S6.
<sup>5</sup> For details of how the categories were formed, please see Table S7.

**Figure 1.**
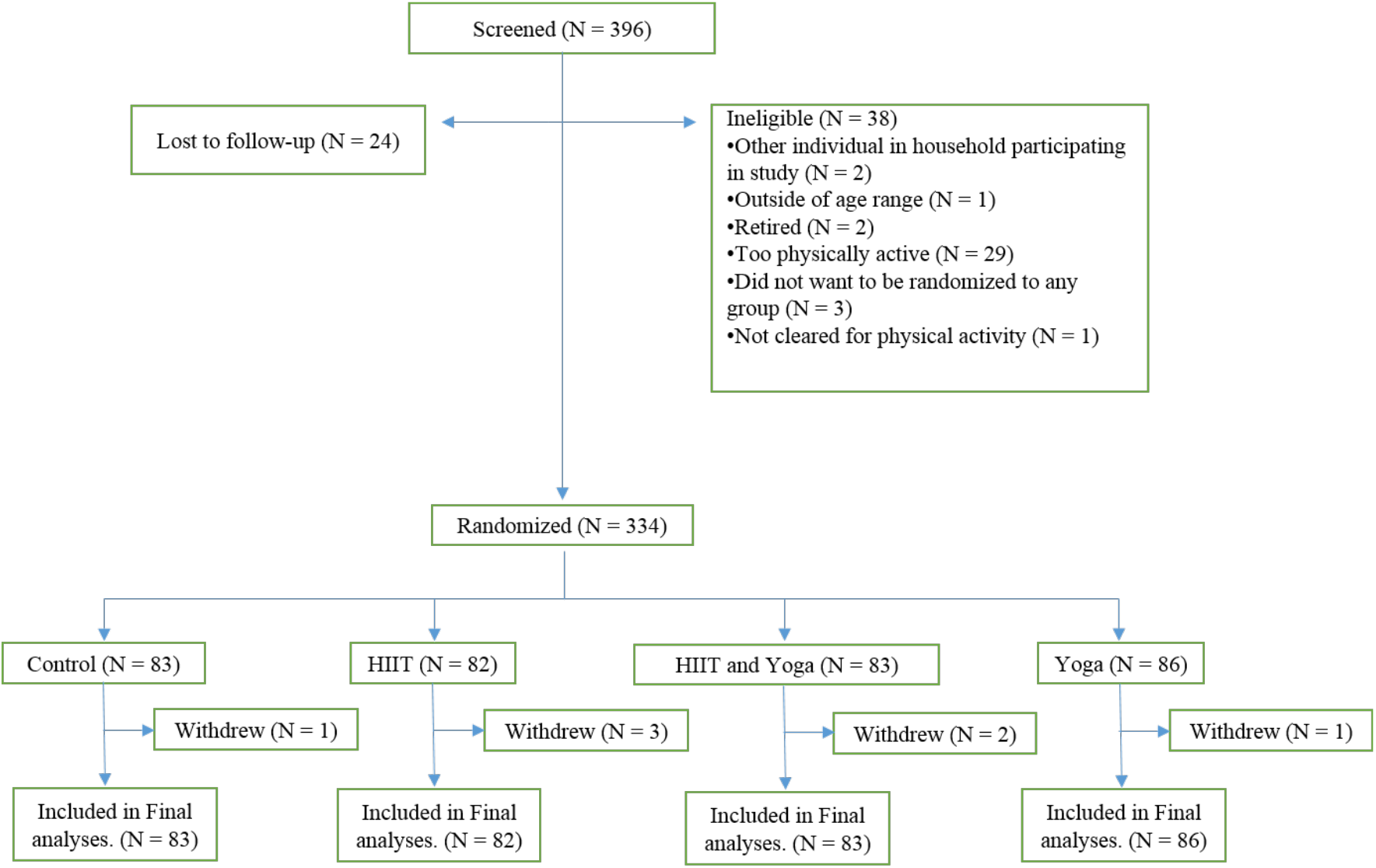
Consort Diagram

### Adherence results

62%, 64%, and 75% of HIIT, Yoga, and HIIT+Yoga participants, respectively, completed 4 or more sessions in the first week of the trial, with an additional 18-29% completing between 1 to 3 workouts (Figure 2). Adherence decreased during the study. While the majority of Yoga and HIIT+Yoga group participants were still meeting the requested four sessions per week by the end of the trial, only 40% in the HIIT group met these activity levels. Weekly completion rate for surveys by group are reported in Table S8.

**Figure 2.**
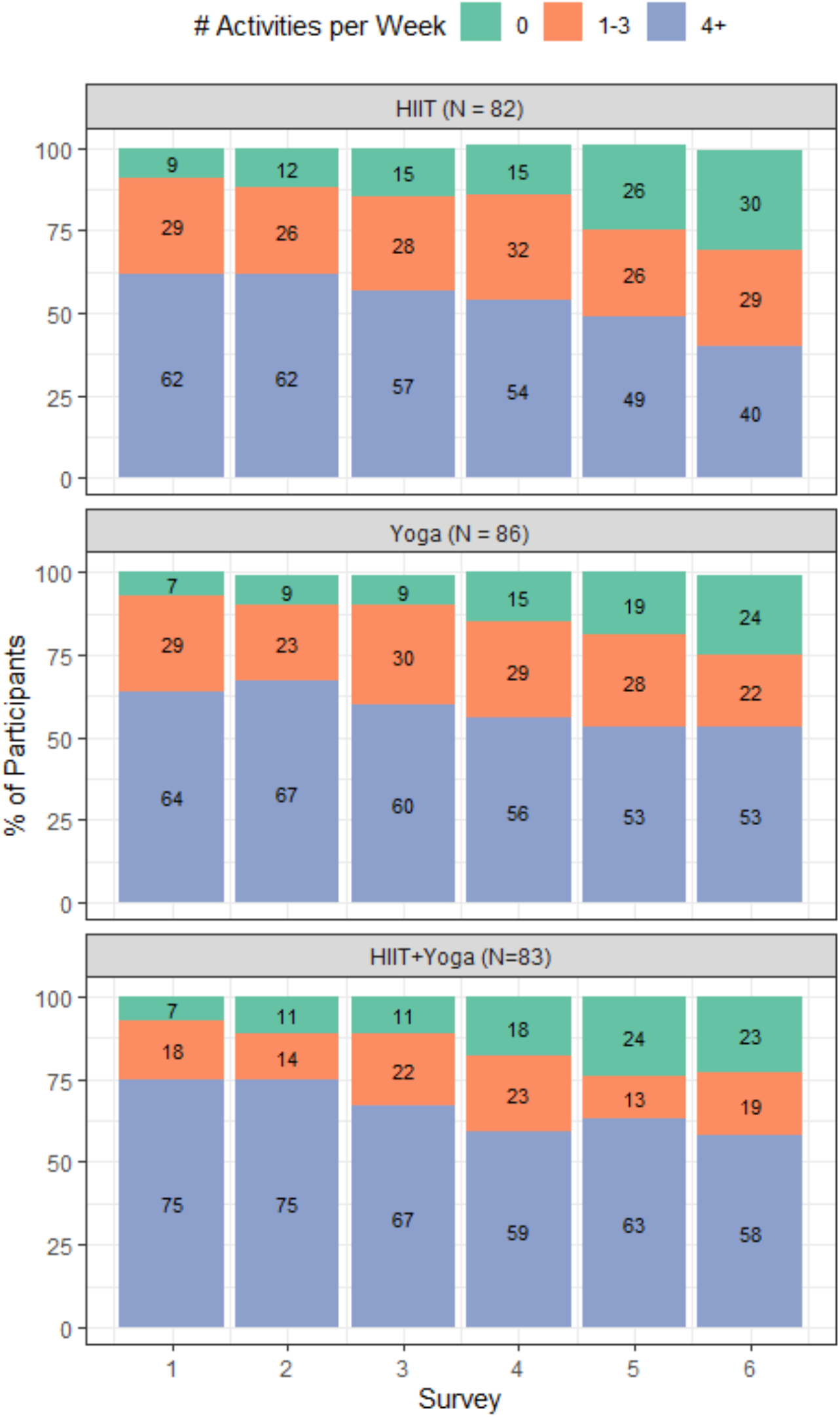
Exercise Adherence Rates by Experimental Condition

### Treatment Groups vs WLC

As seen in Table 2A and Figure 3a, the WLC participants had stable depressive symptoms throughout the 6 weeks (i.e., non-significant S and Q). On the other hand, HIIT and HIIT+Yoga significantly reduced in depressive symptoms over time in non-linear ways (Figure 3a, Table S9 Section A, S10 Section A for HIIT and HIIT+Yoga, respectively), whereas Yoga reduced linearly over time (Figure 3a, Table S11 Section A).

Treatment effect results revealed that baseline estimates for each group were not significantly different from that of the WLC (Table 2, Section B1; all *p*’s > .05), whereas the growth rates over time (specifically the slopes) for HIIT and the HIIT+Yoga were different from WLC (Table 2, Sections B2, B3). At each time point, the ES for Yoga and HIIT+Yoga compared to WLC were significant with small effect sizes (range from -0.12 to -0.31), with ES getting larger over time. In the HIIT group, ES estimates were significant initially, though reduced in size by week 4 when they were no longer significant. See Figure 3c and Table S12 for ES estimates.

**Table 2 –.** Estimates for Trajectories for WLC (A) and Comparisons with Active Treatment Groups (B1-3)

|  | Estimate | SE | 95% CI |
| --- | --- | --- | --- |
| <u>A. Estimates, SE, and 95% CI for intercept (I), slope (S), and quadratic (Q) terms for WLC</u> |  |  |  |
| I | **10.32** | **0.60** | **9.15, 11.49** |
| S | -0.14 | 0.26 | -0.65, 0.38 |
| Q | -0.01 | 0.04 | -0.09, 0.07 |
| <u>B. Estimates for differences between each group and WLC</u> |  |  |  |
| <u>B1. Differences in estimates for I</u> |  |  |  |
| HIIT vs WLC | 0.27 | 0.87 | -1.44, 1.98 |
| Yoga vs WLC | -0.31 | 0.86 | -1.99, 1.37 |
| HIIT+Yoga vs WLC | -1.25 | 0.83 | -2.87, 0.37 |
| <u>B2. Differences in estimates for S</u> |  |  |  |
| HIIT vs WLC | **-0.77** | **0.37** | **-1.50, -0.04** |
| Yoga vs WLC | -0.72 | 0.40 | -1.50, 0.06 |
| HIIT+Yoga vs WLC | **-0.94** | **0.39** | **-1.71, -0.16** |
| <u>B3. Differences in estimates for Q</u> |  |  |  |
| HIIT vs WLC | 0.11 | 0.06 | -0.01, 0.22 |
| Yoga vs WLC | 0.08 | 0.06 | -0.04, 0.20 |
| HIIT+Yoga vs WLC | 0.12 | 0.06 | 0.00, 0.24 |

**Figure 3.**
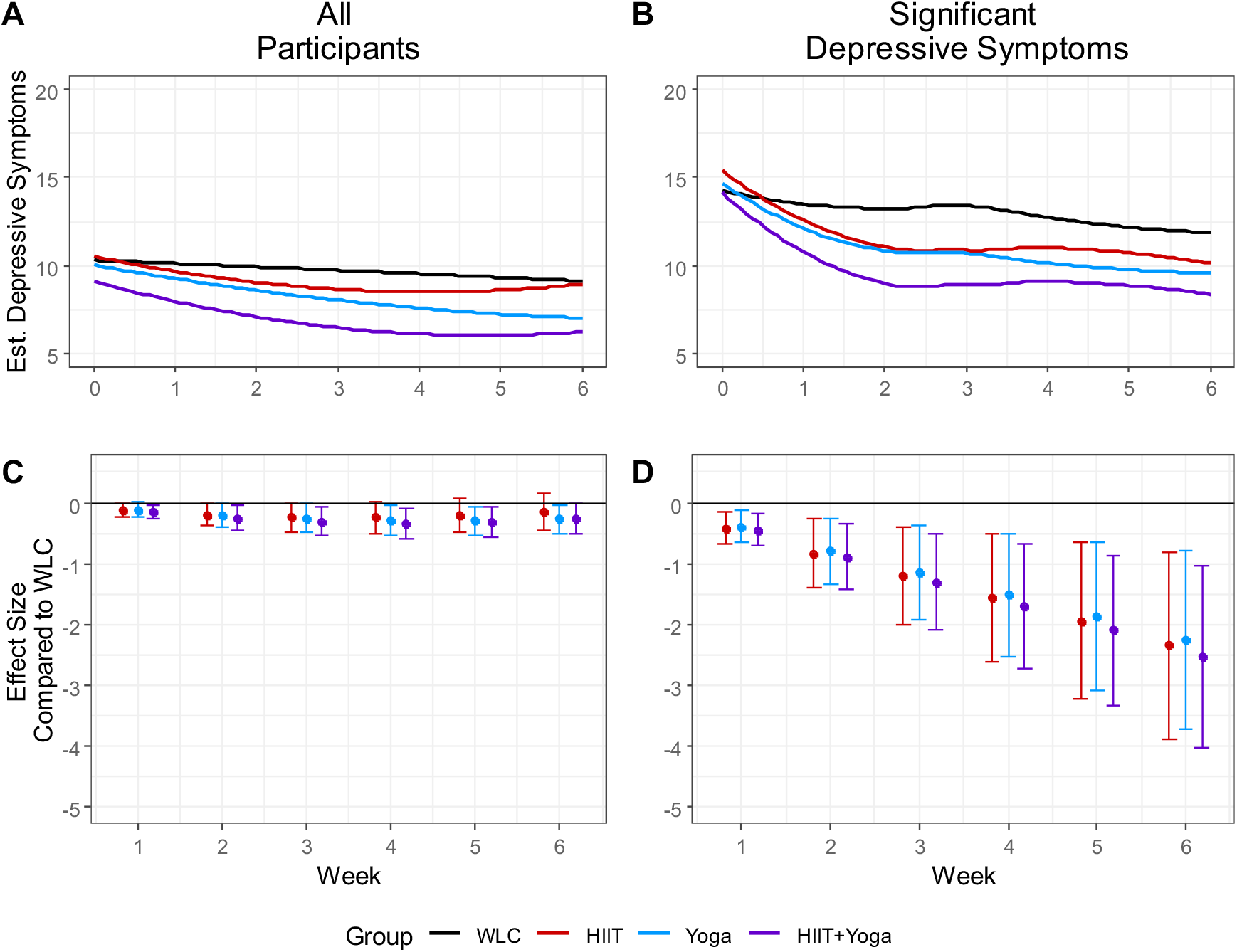
Trajectories and Effect Sizes for Depressive Symptoms Over the Course of the Study Note. Fig 3A shows each group’s trajectories, in the full sample, while figure 3B show the trajectories for those with high (CESD Score ≥ 10) levels of depressive symptoms. Figures 3C-D represent the effect sizes at each time point, for all participants and those with high levels of depressive symptoms, respectively.

### Comparison of Treatment Groups

All three groups had similar trends in decreasing depressive symptoms over the course of the study and effect size estimates for each week were not significantly different from each other (Tables S9-11).

### High Depression Group

When restricted to those participants with pre-randomization baseline ≥10 CESD, (N = 173; Mean = 14.8, SD = 3.98), all treatment groups had significantly greater reductions in depressive symptoms over time compared to the WLC (Table S13, Section B). Rate of decrease in depressive symptoms for each group, in descending strength, was -3.39 (HIIT+Yoga), -3.23 (HIIT), -3.15 (Yoga), and -1.59 (WLC) (See Figure 3b for trends). Within the first week, ES for each treatment group compared to WLC were significant and of small size (ES range -0.40 to -0.45) and continued to grow over the course of the study to very large when the trial was completed (ES range -2.34 to -2.52). See Figure 3d and Table S10 for ES results.

## Discussion

Significant treatment effects in depressive symptoms were observed for participants randomized to complete HIIT or yoga, or a combination of the two, at-home using a suite of mobile applications over a 6-week period compared to WLC participants. While WLC participants’ depressive symptoms remained steady throughout the 6-week period, those in the three active arms had significant declines, as hypothesized. ES at each week were small for all three active groups, with the greatest effects in the combination group. These differences could be attributed to the higher number of HIIT+Yoga participants who completed at least 4 weekly workouts throughout the study. Effects were very large when the sample was restricted to those with high depressive symptoms prior to randomization. Our study reveals an impactful health behavior in which community living adults, especially those with significant depressive symptoms, can engage that can potentially offer relief from the burden of the pandemic.

Prior to the COVID-19 pandemic, percentages of adults across the globe who were already insufficiently active were troublesome,^24^ and evidence suggests that these trends have worsened during the pandemic.^25^ A global call for action^11^ prioritized at-risk groups (e.g. frontline, low-paid, healthcare system workers, laid off adults, older adults) as intervention targets for physical activity programming to reduce risk and severity of infection with COVID-19. Not included in the call for action were those at wider risk for mental health issues, and recent studies are directing attention to individuals in different countries who are at risk for pandemic-related depression. In the UK, for example, 18-34 year old adults, women, and adults with children had the greatest increases in depression during the early months of COVID-19.^5^ Similar elevated risk to women and young adults was identified in the US, as was to those who self-identified as Hispanic, had lower education levels, were not married, or had more life stressors resulting from the pandemic (e.g., financial/employment loss, COVID-19 related death of a family member).^4^ In our study, the majority of participants were women (87%), and nearly half were 18-39 years of age (47%), and had children at home (42%). Yet, a large majority would not be considered to have met significant economic or employment challenges, as many were employed. The extent to which exercise programming can specifically benefit adults with economic challenges remains unclear.

By 2030, the World Economic Forum projected that mental illness will account for US$6 trillion of the annual global economic burden, accounting for more than half the burden from all non-communicable diseases.^26^ With the increasing prevalence of global citizens with COVID-19-related depressive symptoms, the personal, social, and economic burden can be expected to be even more far-reaching and devastating. At the start of the pandemic, exercise was identified as a salient long-term public health target for population-level mental health treatment, even in those not immediately seeking treatment.^1^ The results from the current trial suggest that health officials should continue to promote exercise to the public, directing such promotion especially to those experiencing significant depressive symptoms, with the provision of low-cost or free exercise-based mobile applications to use at home as part of healthcare systems’ initiatives. Free eHealth mobile applications are effective in improving medication adherence and clinic attendance for non-communicable physical diseases, leading to substantial cost-effectiveness.^27^ Perhaps, then, the free provision of exercise-based mobile applications, supported by healthcare systems, could be one method to help reduce the emerging global mental health crisis and its eventual economic burden.

## Supporting information

Consort Checklist

## Data Availability

All data reported here will be available at Open Science Framework (https://osf.io/jbm63/

## Acknowledgments

We would like to acknowledge Anaïs Charbonneau, Abby Cheung, Sarah de Faye, Christina Whang, Carina Wong, Tommy Yang, and Rachel Zhang for their contribution to recruitment and screening of participants, and to Anaïs Charbonneau, Sarah de Faye, and Rachel Zhang for their contribution to data processing.

