## Supplemental File for "The COvid-19 Pandemic and Exercise (COPE) Trial: A multi-group randomized controlled trial comparing effects of an app-based, at-home exercise program to waitlist control on depressive symptoms"

### Supplemental Materials

#### Table of Tables

#### Statistical Approach

##### *Imputation*

Random forest imputation was used to impute weekly Center for Epidemiologic Studies Depression (CESD) item responses when data were missing for one or more responses to the CESD for those participants who had completed some survey data on that given week (Table S1**,** Non-Completely Missing Survey**)**. Imputation was done by taking the data for each week and splitting it into those with surveys and those without. For those with surveys, any items on the CESD with missing data were imputed based on all other participant data from all weeks. Once imputation was complete, the data were rejoined with the data of those without surveys that week. This process was then repeated for each subsequent week. On occasions in which participants did not submit a completed weekly survey (Table S1, Completely Missing Surveys), no imputation was completed since the statistical approach used can handle missing data.

#### Demographic Data Coding

Several of the demographic variables were group together for ease of communication in tables. This was done for income (Table S3), employment (Table S4), education (Table S5), cultural background (Table S6) and marital status (Table S7).

**Mplus Code**

***All Participants Sample, Including Treatment Groups***

```
  VARIABLE: NAMES ARE id cond g1 g2 g3 male age
                CESD0 CESD1 CESD2 CESD3 CESD4 CESD5 CESD6 CESD12;
      MISSING=ALL(999);
      USEVARIABLES ARE g1 g2 g3
                CESD0 CESD1 CESD2 CESD3 CESD4 CESD5 CESD6;
   ANALYSIS: ESTIMATOR = MLR;
           STITERATIONS=20000;
           ITERATION = 40000;

    MODEL:
        i s q |  CESD0@0  CESD1@1  CESD2@2 CESD3@3 CESD4@4 CESD5@5 CESD6@6;
        i s q ON g1 g2 g3;

        s ON g1 (bs1);
        s ON g2 (bs2);
        s ON g3 (bs3);
        q ON g1 (bq1);
        q ON g2 (bq2);
        q ON g3 (bq3);
        i(v0);
        CESD0 - CESD6 (r0-r6);

MODEL CONSTRAINT:
       NEW(g1dt1 g1dt2 g1dt3 g1dt4 g1dt5 g1dt6);
       g1dt1 = (bs1*1 + bq1*1)/sqrt(v0+r0/2+r1/2);
       g1dt2 = (bs1*2 + bq1*4)/sqrt(v0+r0/3+r1/3+r2/3);
       g1dt3 = (bs1*3 + bq1*9)/sqrt(v0+r0/4+r1/4+r2/4+r3/4);
       g1dt4 = (bs1*4 + bq1*16)/sqrt(v0+r0/5+r1/5+r2/5+r3/5+r4/5);
       g1dt5 = (bs1*5 + bq1*25)/sqrt(v0+r0/6+r1/6+r2/6+r3/5 +r4/6+r5/6);
       g1dt6 = (bs1*6 + bq1*36)/sqrt(v0+r0/7+r1/7+r2/7+r3/7 +r4/7+r5/7+r6/7);

       NEW(g2dt1 g2dt2 g2dt3 g2dt4 g2dt5 g2dt6);
       g2dt1 = (bs2*1 + bq2*1)/sqrt(v0+r0/2+r1/2);
       g2dt2 = (bs2*2 + bq2*4)/sqrt(v0+r0/3+r1/3+r2/3);
       g2dt3 = (bs2*3 + bq2*9)/sqrt(v0+r0/4+r1/4+r2/4+r3/4);
       g2dt4 = (bs2*4 + bq2*16)/sqrt(v0+r0/5+r1/5+r2/5+r3/5+r4/5);
       g2dt5 = (bs2*5 + bq2*25)/sqrt(v0+r0/6+r1/6+r2/6+r3/5+r4/6+r5/6);
       g2dt6 = (bs2*6 + bq2*36)/sqrt(v0+r0/7+r1/7+r2/7+r3/7+r4/7+r5/7+r6/7);

      NEW(g3dt1 g3dt2 g3dt3 g3dt4 g3dt5 g3dt6);
       g3dt1 = (bs3*1 + bq3*1)/sqrt(v0+r0/2+r1/2);
       g3dt2 = (bs3*2 + bq3*4)/sqrt(v0+r0/3+r1/3+r2/3);
       g3dt3 = (bs3*3 + bq3*9)/sqrt(v0+r0/4+r1/4+r2/4+r3/4);
       g3dt4 = (bs3*4 + bq3*16)/sqrt(v0+r0/5+r1/5+r2/5+r3/5+r4/5);
```

```
    g3dt5 = (bs3*5 + bq3*25)/sqrt(v0+r0/6+r1/6+r2/6+r3/5+r4/6+r5/6);
    g3dt6 = (bs3*6 + bq3*36)/sqrt(v0+r0/7+r1/7+r2/7+r3/7+r4/7+r5/7+r6/7);

OUTPUT: SAMPSTAT CINTERVAL STANDARDIZED RESIDUAL MODINDICES (3.84);
```

***High Depression Sample, Including Treatment Groups – Free Time Scores***

```
VARIABLE: NAMES ARE id cond g1 g2 g3 male age
              CESD0 CESD1 CESD2 CESD3 CESD4 CESD5 CESD6 CESD12;

  MISSING=ALL(999);

  USEVARIABLES ARE g1 g2 g3 CESD0 CESD1 CESD2 CESD3 CESD4 CESD5 CESD6;
 ANALYSIS: ESTIMATOR = MLR;
       STITERATIONS=5000;
       ITERATION = 20000;

 MODEL:
   i s | CESD0@0  CESD1@1  CESD2* CESD3* CESD4* CESD5* CESD6*;

   i s ON g1 g2 g3;
   s ON g1 (bs1);
   s ON g2 (bs2);
   s ON g3 (bs3);
   i(v0);
   CESD1 - CESD6 (r1-r6);
   CESD0@0;
   CESD1 WITH CESD2-CESD4;
   CESD2 WITH CESD3-CESD5;
   CESD3 WITH CESD4-CESD6;
   CESD4 WITH CESD5-CESD6;
   CESD5 WITH CESD6;

 MODEL CONSTRAINT:
   NEW(g1dt1 g1dt2 g1dt3 g1dt4 g1dt5 g1dt6);
   g1dt1 = (bs1*1)/sqrt(v0+r1);
   g1dt2 = (bs1*2)/sqrt(v0+r1/2+r2/2);
   g1dt3 = (bs1*3)/sqrt(v0+r1/3+r2/3+r3/3);
   g1dt4 = (bs1*4)/sqrt(v0+r1/4+r2/4+r3/4+r4/4);
   g1dt5 = (bs1*5)/sqrt(v0+r1/5+r2/5+r3/5+r4/5+r5/5);
   g1dt6 = (bs1*6)/sqrt(v0+r1/6+r2/6+r3/6+r4/6+r5/6+r6/6);

  NEW(g2dt1 g2dt2 g2dt3 g2dt4 g2dt5 g2dt6);
   g2dt1 = (bs2*1)/sqrt(v0+r1);
   g2dt2 = (bs2*2)/sqrt(v0+r1/2+r2/2);
   g2dt3 = (bs2*3)/sqrt(v0+r1/3+r2/3+r3/3);
   g2dt4 = (bs2*4)/sqrt(v0+r1/4+r2/4+r3/4+r4/4);
   g2dt5 = (bs2*5)/sqrt(v0+r1/5+r2/5+r3/5+r4/5+r5/5);
   g2dt6 = (bs2*6)/sqrt(v0+r1/6+r2/6+r3/6+r4/6+r5/6+r6/6);

  NEW(g3dt1 g3dt2 g3dt3 g3dt4 g3dt5 g3dt6);
   g3dt1 = (bs3*1)/sqrt(v0+r1);
```

```
    g3dt2 = (bs3*2)/sqrt(v0+r1/2+r2/2);
    g3dt3 = (bs3*3)/sqrt(v0+r1/3+r2/3+r3/3);
    g3dt4 = (bs3*4)/sqrt(v0+r1/4+r2/4+r3/4+r4/4);
    g3dt5 = (bs3*5)/sqrt(v0+r1/5+r2/5+r3/5+r4/5+r5/5);
    g3dt6 = (bs3*6)/sqrt(v0+r1/6+r2/6+r3/6+r4/6+r5/6+r6/6);

OUTPUT: SAMPSTAT CINTERVAL STANDARDIZED RESIDUAL;
```

*Table S1 - Missing Surveys at Item and Survey Levels.*

| Week | Non-missing Survey (All items complete) | Non-Completely Missing Survey (Missing >= 1 items) | Completely Missing Surveys |
|---|---|---|---|
| 0 | 327 | 7 | 0 |
| 1 | 302 | 9 | 23 |
| 2 | 299 | 5 | 30 |
| 3 | 290 | 2 | 42 |
| 4 | 287 | 6 | 41 |
| 5 | 276 | 6 | 52 |
| 6 | 280 | 6 | 48 |

*Table S2 - Fit Indices for the Primary SEM Models.*

| **Measure** | **All Participants - quadratic time** | **High Depression – Free Time Scores** |
|---|---|---|
| Root Mean Square Error Of Approximation, Est. (.90CI) | 0.047 (0.025, 0.067) | 0.043 (0.000, 0.081) |
| CFI | 0.984 | 0.990 |
| Standardized Root Mean Square Residual | 0.024 | 0.041 |

*Table S3 - Income Grouping.*

| **Income Values** | **Recoded** |
|---|---|
| 1 - 10,000<br>10,001 - 20,000<br>20,001 - 30,000<br>30,001 - 40,000 | 0 – 40,000 |
| 40,001 - 50,000,<br>50,001 - 60,000,<br>60,001 - 70,000,<br>70,001 - 80,000 | 40,001 – 80,000 |
| 80,001 - 90,000,<br>90,001 - 100,000,<br>100,001 - 110,000,<br>110,001 - 120,000 | 80,001 – 120,000 |
| 120,001 - 130,000,<br>130,001 - 140,000,<br>140,001 - 150,000,<br>150,001 - 160,000 | 120,001 – 160,000 |
| 160,001 - 170,000,<br>170,001 - 180,000,<br>180,001 - 190,000,<br>190,001 - 200,000,<br>200,001 - 210,000,<br>210,001 - 220,000, | 160,000 + |

| | |
|---|---|
| 220,001 - 230,000,<br><br>230,001 - 240,000,<br><br>240,001 - 250,000,<br><br>250,001+ | |
| Do not know / prefer not to answer,<br><br>[Missing] | No response |

*Table S4 - Employment Grouping.*

| Current Employment | Other (Text Response) | Coded Value |
|---|---|---|
| Other | Full time work with one furlough day due to covid | Full Time |
| Other | teacher in spring semester | Full Time |
| Other | Teacher summer vacation | Full Time |
| Working full-time | | Full Time |
| Working full-time,Other | on summer vacation | Full Time |
| Working full-time,Self-employed | | Full Time |
| Working full-time,Self-employed ,Other | I'm working full time at a company, but also self-employed teaching (Not full-time-self-employed) | Full Time |
| Homemaker | | Not Working |
| Looking for work; unemployed | | Not Working |
| Looking for work; unemployed,Homemaker | | Not Working |
| Looking for work; unemployed,Other | housewife | Not Working |
| Looking for work; unemployed,Other | working - unpaid | Not Working |
| Looking for work; unemployed,Temporarily laid off | | Not Working |
| Maternity or sick leave (volunteered) | | Not Working |
| Other | Leave of absence | Not Working |

| | | |
|---|---|---|
| Other | Self employed but no work available | Not Working |
| Other | Stay at home parent | Not Working |
| Self-employed ,Looking for work; unemployed,Permanently disabled (volunteered),Other | Home based business not in market due to covid | Not Working |
| Self-employed ,Temporarily laid off | | Not Working |
| Temporarily laid off | | Not Working |
| Temporarily laid off,Maternity or sick leave (volunteered) | | Not Working |
| [Missing] | | Other |
| Don't know/not sure | | Other |
| Other | Will be laid off during this study | Other |
| Other | | Other |
| Prefer not to say | | Other |
| Self-employed | | Other |
| Working part-time | | Part Time |
| Working part-time,Homemaker | | Part Time |
| Working part-time,Other | on CEWS | Part Time |
| Working part-time,Self-employed | | Part Time |
| Working part-time,Self-employed ,Homemaker | | Part Time |
| Full-time student | | Student |

| | | |
|---|---|---|
| Looking for work; unemployed,Full-time student | | Student |
| Looking for work; unemployed,Part-time student | | Student |
| Looking for work; unemployed,Temporarily laid off,Part-time student | | Student |
| Part-time student | | Student |
| Self-employed ,Full-time student | | Student |
| Temporarily laid off,Full-time student | | Student |
| Temporarily laid off,Part-time student | | Student |
| Working full-time,Full-time student | | Student |
| Working full-time,Part-time student | | Student |
| Working part-time,Full-time student | | Student |
| Working part-time,Full-time student,Other | Support from parents | Student |
| Working part-time,Part-time student | | Student |

*Table S5 - Education Grouping.*

| **Educational Status** | **Recoded** |
|---|---|
| High school diploma or a high school equivalency certificate | High School or less |
| College, CEGEP or other non-university certificate or diploma (other than trades certificates or diplomas), | College, trade school or certificate |
| Trade certificate or diploma, | College, trade school or certificate |
| University certificate or diploma below the bachelor's level | College, trade school or certificate |
| Bachelor's degree (e.g. B.A., B.Sc., LL.B.) | Bachelor or equivalent |
| University certificate, diploma, degree above the bachelor's level | Postgraduate / professional training |
| Prefer not to answer, | No response |
| [Missing] | No response |

*Note: "Less than high school diploma or its equivalent" was an option, but no participant selected it.*

*Table S6 - Cultural Background Grouping.*

| **Cultural Background** | **Other (Text Response)** | **Coded Value** |
| --- | --- | --- |
| Chinese | | Asian |
| Chinese,Filipino | | Asian |
| Chinese,South East Asian (e.g., Vietnamese, Cambodian, Malaysian, Laotian, etc) | | Asian |
| Filipino | | Asian |
| Filipino,South East Asian (e.g., Vietnamese, Cambodian, Malaysian, Laotian, etc) | | Asian |
| Japanese | | Asian |
| Korean | | Asian |
| Other - please specify | Taiwanese | Asian |
| South Asian (e.g., East Indian, Pakistani, Sri Lankan, etc) | | Asian |
| South East Asian (e.g., Vietnamese, Cambodian, Malaysian, Laotian, etc) | | Asian |
| Aboriginal decent (e.g., North American Indian, Métis or Inuit (Eskimo)) | | Other |
| Aboriginal decent (e.g., North American Indian, Métis or Inuit (Eskimo)),Black (e.g., African, Haitian, Jamaican, Somali, etc) | | Other |
| Aboriginal decent (e.g., North American Indian, Métis or Inuit (Eskimo)),White | | Other |

| | | |
|---|---|---|
| Arab | | Other |
| Black (e.g., African, Haitian, Jamaican, Somali, etc) | | Other |
| Latin American | | Other |
| Other - please specify | | Other |
| Other - please specify | european / mixed | Other |
| Other - please specify | Doesn't let you choose more than one. Aboriginal and European. | Other |
| Other - please specify | Afghan | Other |
| Other - please specify | Austrian | Other |
| South Asian (e.g., East Indian, Pakistani, Sri Lankan, etc),Latin American | | Other |
| West Asian (e.g., Iranian, Afghan, etc) | | Other |
| White,Arab | | Other |
| White,Chinese | | Other |
| White,Chinese,South East Asian (e.g., Vietnamese, Cambodian, Malaysian, Laotian, etc) | | Other |
| White,Filipino | | Other |
| White,Japanese | | Other |
| White,Latin American | | Other |
| White,Other - please specify | Jewish | Other |

| | | |
|---|---|---|
| White,Other - please specify | European | Other |
| White,Other - please specify | Ashkenazi Jewish | Other |
| White,Other - please specify | Scandinavian | Other |
| White,Other - please specify | Dutch | Other |
| White,South East Asian (e.g., Vietnamese, Cambodian, Malaysian, Laotian, etc) | | Other |
| Other - please specify | Italian | White |
| Other - please specify | IRISH | White |
| White | | White |

*Note: As cultural backgrounds were asked as check boxes, many participant selected multiple options, making it difficult to define groups. Therefore, participants were split into White, Asian and Other, as White and Asian participants made up a large majority of the sample.*

*Table S7 - Marital Status Groupings.*

| **Marital Status** | **Recoded** |
|---|---|
| Married | Married |
| Living common-law | Married |
| Separated | No Longer Married |
| Divorced | No Longer Married |
| Widowed | No Longer Married |
| Single, never married | Single |
| Prefer not to answer | Other |
| [Missing] | Other |

*Table S8 - Weekly Survey Response Rate by Experimental Grouping.*

| | Week | | | | | | |
|---|---|---|---|---|---|---|---|
| | **0** | **1** | **2** | **3** | **4** | **5** | **6** |
| WLC | 83 (100%) | 78 (94%) | 77 (93%) | 80 (96%) | 76 (92%) | 77 (93%) | 74 (89%) |
| HIIT | 82 (100%) | 77 (94%) | 76 (93%) | 69 (84%) | 72 (88%) | 64 (78%) | 70 (85%) |
| Yoga | 86 (100%) | 81 (94%) | 76 (88%) | 74 (86%) | 76 (88%) | 75 (87%) | 74 (86%) |
| HIIT+Yoga | 83 (100%) | 75 (90%) | 75 (90%) | 69 (83%) | 69 (83%) | 66 (80%) | 68 (82%) |

*Table S9 - Estimates for Trajectories for HIIT (A) and Comparisons with WLC, Yoga and HIIT+Yoga Groups (B1-3).*

| | **Estimate** | **SE** | **95% CI** |
|---|---|---|---|
| A. Estimates, SE, and 95% CI for intercept (I; estimated baseline), slope (S; time), and quadratic (Q; time$^2$) terms for HIIT | | | |
| I | **10.59** | **0.64** | **9.33, 11.85** |
| S | **-0.91** | **0.27** | **-1.43, -0.38** |
| Q | **0.10** | **0.04** | **0.02, 0.17** |
| B. Estimates for differences between each group and WLC | | | |
| *B1. Differences in estimates for I* | | | |
| WLC vs HIIT | -0.27 | 0.87 | -1.98, 1.44 |
| Yoga vs HIIT | -0.58 | 0.89 | -2.33, 1.17 |
| HIIT+Yoga vs HIIT | -1.52 | 0.87 | -3.22, 0.18 |
| *B2. Differences in estimates for S* | | | |
| WLC vs HIIT | **0.77** | **0.37** | **0.04, 1.50** |
| Yoga vs HIIT | 0.05 | 0.4 | -0.74, 0.84 |
| HIIT+Yoga vs HIIT | -0.16 | 0.4 | -095% , 0.62 |
| *B3. Differences in estimates for Q* | | | |
| WLC vs HIIT | -0.11 | 0.06 | -0.22, 0.01 |
| Yoga vs HIIT | -0.03 | 0.06 | -0.14, 0.09 |
| HIIT+Yoga vs HIIT | 0.01 | 0.06 | -0.11, 0.13 |

*Note: Results from the SEM. model estimating intercept, slope, and quadratic term for HIIT group (Section A) and comparisons of these estimates with those of the three other groups (WLC, Yoga, HIIT+Yoga; Section B). Bold text denotes $p < .05$*

*Table S10 - Estimates for Trajectories for HIIT+Yoga (A) and Comparisons with WLC, HIIT and Yoga Groups (B1-3).*

| | **Estimate** | **SE** | **95% CI** |
|---|---|---|---|
| A. Estimates, SE, and 95% CI for intercept (I; estimated baseline), slope (S; time), and quadratic (Q; time$^2$) terms for HIIT+Yoga | | | |
| I | **9.07** | **0.58** | **7.94, 10.21** |
| S | **-1.07** | **0.30** | **-1.66, -0.49** |
| Q | **0.11** | **0.05** | **0.02, 0.20** |
| B. Estimates for differences between each group and WLC | | | |
| *B1. Differences in estimates for I* | | | |
| WLC vs HIIT+Yoga | 1.25 | 0.83 | -0.37, 2.87 |
| HIIT vs HIIT+Yoga | 1.52 | 0.87 | -0.18, 3.22 |
| Yoga vs HIIT+Yoga | 0.94 | 0.85 | -0.72, 2.60 |
| *B2. Differences in estimates for S* | | | |
| WLC vs HIIT+Yoga | **0.94** | **0.39** | **0.16, 1.71** |
| HIIT vs HIIT+Yoga | 0.17 | 0.40 | -0.62, 095% |
| Yoga vs HIIT+Yoga | 0.21 | 0.42 | -0.62, 1.04 |
| *B3. Differences in estimates for Q* | | | |
| WLC vs HIIT+Yoga | -0.12 | 0.06 | -0.24, 0.00 |
| HIIT vs HIIT+Yoga | -0.01 | 0.06 | -0.13, 0.11 |
| Yoga vs HIIT+Yoga | -0.04 | 0.06 | -0.16, 0.09 |

*Note: Results from the SEM model estimating intercept, slope, and quadratic term for HIIT+Yoga group (Section A) and comparisons of these estimates with those of the three other groups (WLC, HIIT, Yoga; Section B). Bold text denotes p < .05*

*Table S11 - Estimates for Trajectories for Yoga (A) and Comparisons with WLC, HIIT and HIIT+Yoga Groups (B1-3).*

| | **Estimate** | **SE** | **95% CI** |
|---|---|---|---|
| A. Estimates, SE, and 95% CI for intercept (I; estimated baseline), slope (S; time), and quadratic (Q; time$^2$) terms for Yoga | | | |
| I | **10.01** | **0.62** | **8.79, 11.23** |
| S | **-0.86** | **0.31** | **-1.46, -0.26** |
| Q | 0.07 | 0.05 | -0.02, 0.16 |
| B. Estimates for differences between each group and WLC | | | |
| *B1. Differences in estimates for I* | | | |
| WLC vs Yoga | 0.31 | 0.86 | -1.37, 1.99 |
| HIIT vs Yoga | 0.58 | 0.89 | -1.17, 2.33 |
| HIIT+Yoga vs Yoga | -0.94 | 0.85 | -2.60, 0.72 |
| *B2. Differences in estimates for S* | | | |
| WLC vs Yoga | 0.72 | 0.40 | -0.06, 1.50 |
| HIIT vs Yoga | -0.05 | 0.40 | -0.84, 0.74 |
| HIIT+Yoga vs Yoga | -0.21 | 0.422 | -1.04, 0.62 |
| *B3. Differences in estimates for Q* | | | |
| WLC vs Yoga | -0.08 | 0.06 | -0.20, 0.04 |
| HIIT vs Yoga | 0.03 | 0.06 | -0.09, 0.14 |
| HIIT+Yoga vs Yoga | 0.04 | 0.06 | -0.09, 0.16 |

*Note: Results from the SEM. model estimating intercept, slope, and quadratic term for HIIT group (Section A) and comparisons of these estimates with those of the three other groups (WLC, HIIT, HIIT+Yoga; Section B). Bold text denotes p < .05*

*Table 12 - Effect Sizes for Model with all Individuals and Model including only those with High Depressive Symptoms at Baseline.*

**All Participants**

| **Time** | **HIIT** | | **Yoga** | | **HIIT+Yoga** | |
|---|---|---|---|---|---|---|
| | **Effect Size** | **95% CI** | **Effect Size** | **95% CI** | **Effect Size** | **95% CI** |
| Week 1 | -0.12 | [-0.23, -0.01] | -0.11 | [-0.23, 0.00] | -0.14 | [-0.26, -0.03] |
| Week 2 | -0.20 | [-0.39, -0.01] | -0.20 | [-0.39, 0.00] | -0.25 | [-0.44, -0.05] |
| Week 3 | -0.24 | [-0.48, 0.00] | -0.26 | [-0.50, -0.02] | -0.31 | [-0.55, -0.07] |
| Week 4 | -0.25 | [-0.50, 0.01] | -0.29 | [-0.53, -0.05] | -0.33 | [-0.58, -0.08] |
| Week 5 | -0.21 | [-0.48, 0.06] | -0.29 | [-0.53, -0.06] | -0.31 | [-0.56, -0.07] |
| Week 6 | -0.15 | [-0.45, 0.16] | -0.27 | [-0.51, -0.04] | -0.26 | [-0.51, 0.00] |

**Subpopulation with High Depressive Symptoms**

| **Time** | **HIIT** | | **Yoga** | | **HIIT+Yoga** | |
|---|---|---|---|---|---|---|
| | **Effect Size** | **95% CI** | **Effect Size** | **95% CI** | **Effect Size** | **95% CI** |
| Week 1 | -0.41 | [-0.69, -0.14] | -0.40 | [-0.66, -0.14] | -0.45 | [-0.71, -0.18] |
| Week 2 | -0.83 | [-1.39, -0.27] | -0.80 | [-1.33, -0.27] | -0.90 | [-1.44, -0.36] |
| Week 3 | -1.20 | [-2.01, -0.40] | -1.16 | [-1.93, -0.38] | -1.30 | [-2.08, -0.52] |

| | | | | | | |
|---|---|---|---|---|---|---|
| Week 4 | -1.57 | [-2.62, -0.52] | -1.51 | [-2.52, -0.50] | -1.70 | [-2.71, -0.68] |
| Week 5 | -1.94 | [-3.22, -0.66] | -1.86 | [-3.09, -0.64] | -2.09 | [-3.33, -0.86] |
| Week 6 | -2.34 | [-3.87, -0.80] | -2.25 | [-3.71, -0.78] | -2.52 | [-4.01, -1.04] |

*Note: All effect sizes are compared to the WLC group.*

*Table 13 - Estimates for Trajectories for WLC (A) and Comparisons with WLC, HIIT and HIIT+Yoga Groups (B1 & B2) in Participants with High Levels of Depression Symptoms at Baseline.*

| | **Estimate** | **SE** | **95% CI** |
|---|---|---|---|
| A. Estimates, SE, and 95% CI for intercept (I) and slope (S) terms for WLC | | | |
| I | **14.41** | **0.56** | **13.32, 15.51** |
| S | **-1.16** | **0.45** | **-2.03, -0.29** |
| B. Estimates for differences between each group and WLC | | | |
| *B1. Differences in estimates for I* | | | |
| HIIT vs WLC | 0.982 | 0.86 | -0.70, 2.66 |
| Yoga vs WLC | 0.520 | 0.81 | -1.06, 2.10 |
| HIIT+Yoga vs WLC | -0.26 | 0.83 | -1.89, 1.38 |
| *B2. Differences in estimates for S* | | | |
| HIIT vs WLC | **-2.07** | **0.68** | **-3.41, -0.73** |
| Yoga vs WLC | **-1.99** | **0.65** | **-3.27, -0.71** |
| HIIT+Yoga vs WLC | **-2.24** | **0.67** | **-3.54, -0.93** |

*Note: Results from the SEM model, including only those with high levels of depressive symptoms at baseline, estimating intercept and slope for waitlist control (Section A) and comparisons of these estimates with those of the three active groups (HIIT, Yoga, HIIT+Yoga; Section B). Bold text denotes p < .05*

**Figures**

*Figure S1 - SEM Path Diagram for Model including all Participants.*

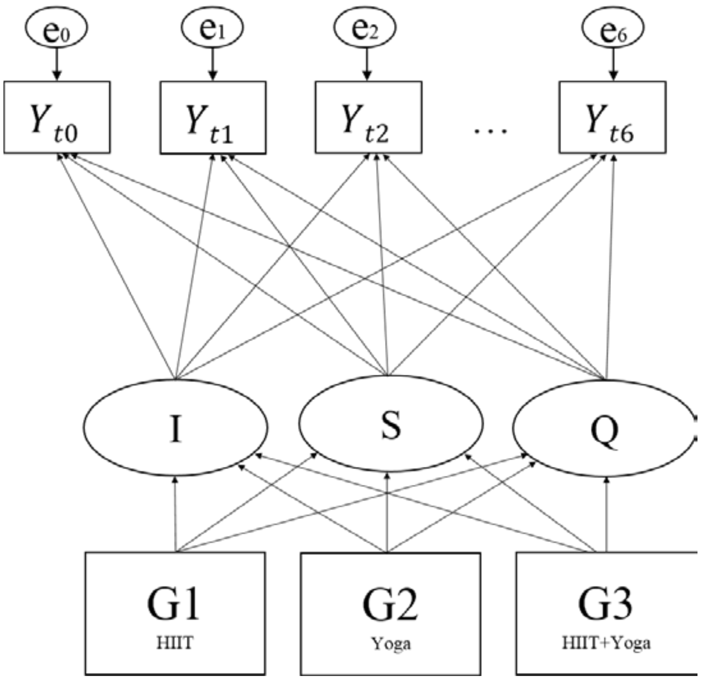

*Figure S2 - SEM Path Diagram for Model including Participants with High Baseline Depressive Symptoms.*

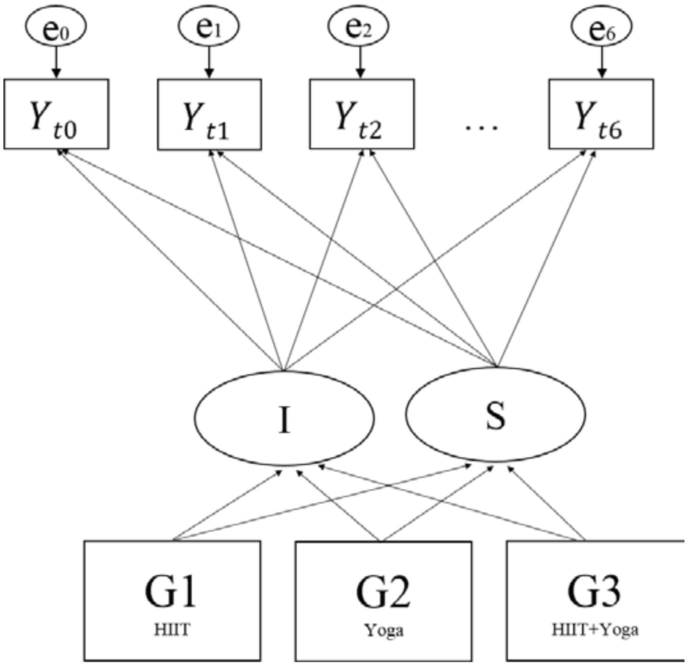
